# Interpretation of wide confidence intervals in meta-analytic estimates: Is the ‘Absence of Evidence’ ‘Evidence of Absence’?

**DOI:** 10.1101/2023.07.11.23292513

**Authors:** Sarah L. Miller, Jordan Tuia, Vinay Prasad

## Abstract

**Introduction:** Recently, a Cochrane review by Jefferson et al. on physical interventions to slow the spread of respiratory viruses concluded that, “Wearing masks in the community probably makes little or no difference to the outcome of laboratory-confirmed influenza/SARS-CoV-2 compared to not wearing masks”, though this finding had a wide confidence interval. Cochrane issued a rare clarifying statement, fueling controversy. We sought to contextualize the findings of the review by Jefferson et al.

**Methods:** We searched for consecutive reviews by Cochrane published on or before March 9th, 2023. We included studies where a central finding showed an intervention offered no statistically significant benefit, and ascertained the language used by reviewers to describe that result. We compare this to the report by Jefferson et al., and deemed it consistent or inconsistent with the language of their report.

**Results:** We found between November 21^st^, 2022, and March 9^th^, 2023, there were 20 Cochrane reviews that met the inclusion criteria. We found that 95% (n = 19) of the reviews used language that was consistent with Jefferson’s findings, while 5% (n = 1) used language inconsistent with Jefferson’s conclusion, describing the effect of the intervention on the outcome as “unclear”.

**Discussion:** Most reviews performed by Cochrane conclude that interventions which fail to show statistically significant benefits make “no difference” have “no effect” or do not “increase or decrease” the outcome, and this occurs despite wide confidence intervals. The conclusions by Jefferson et al. are consistent with Cochrane reporting guidelines and clarification from the organization was unjustified.

## Introduction

An updated Cochrane review on physical interventions to slow the spread of respiratory viruses^1^ has sparked debate among researchers and in the media over the interpretation of the results, leading Cochrane’s editor-in-chief to issue a statement attempting to clarify comments made by the lead author^2^.

Among other topics, the review examined the effect of medical or surgical masks on the spread of respiratory viruses in the community and found a relative risk of 1.01 95% CI (0.72 - 1.42) after pooling 6 trials. The authors of the Cochrane review concluded, “Wearing masks in the community probably makes little or no difference to the outcome of laboratory-confirmed influenza/SARS-CoV-2 compared to not wearing masks’’, and the first author and senior reviewer Tom Jefferson was quoted by a news outlet saying, “There is just no evidence that they make any difference. Full stop.”^3^

In response, Cochrane editor-in-chief Karla Soares-Weiser, issued an unprecedented clarification, stating, “Many commentators have claimed that a recently-updated Cochrane Review shows that ‘masks don’t work’, which is an inaccurate and misleading interpretation. It would be accurate to say that the review examined whether interventions to promote mask wearing help to slow the spread of respiratory viruses, and that the results were inconclusive.” The editor went on to specifically criticize Jefferson, “Soares-Weiser also said, though, that one of the lead authors of the review even more seriously misinterpreted its finding on masks by saying in an interview that it proved ‘there is just no evidence that they make any difference.’ In fact, Soares-Weiser said, ‘that statement is not an accurate representation of what the review found.’”^4^

Dueling interpretations of the central findings beg the question of consistency. In the case of masking to slow the spread of respiratory viruses, masking proponents point to a wide confidence interval, which includes values compatible with moderate benefit (e.g., 0.8) as evidence that the analysis cannot show ‘masks don’t work’ but rather ‘masks might work, with longer or better studies.’ In contrast, critics point to the fact that, as a general rule in medicine, interventions that fail to show benefit despite repeated testing are considered negative interventions. As such we sought to empirically assess the interpretation of Cochrane reviews with broad confidence intervals spanning one.

## Methods

### Data collection

We searched the Cochrane Library for reviews where a central finding had a confidence interval that crossed one. We included studies that were: 1. Intervention type reviews 2. Had a main outcome measured as a risk ratio, odds ratio, or hazard ratio 3. Had a confidence interval spanning one with moderate or high certainty evidence. 4. Published on or before March 9^th^, 2023. One reviewer (SM) selected twenty consecutive studies that met the inclusion criteria. We extracted quotes regarding Cochrane’s interpretation listed in the **Table**. The **Figure** shows the corresponding point estimates and confidence intervals.

**Table.**
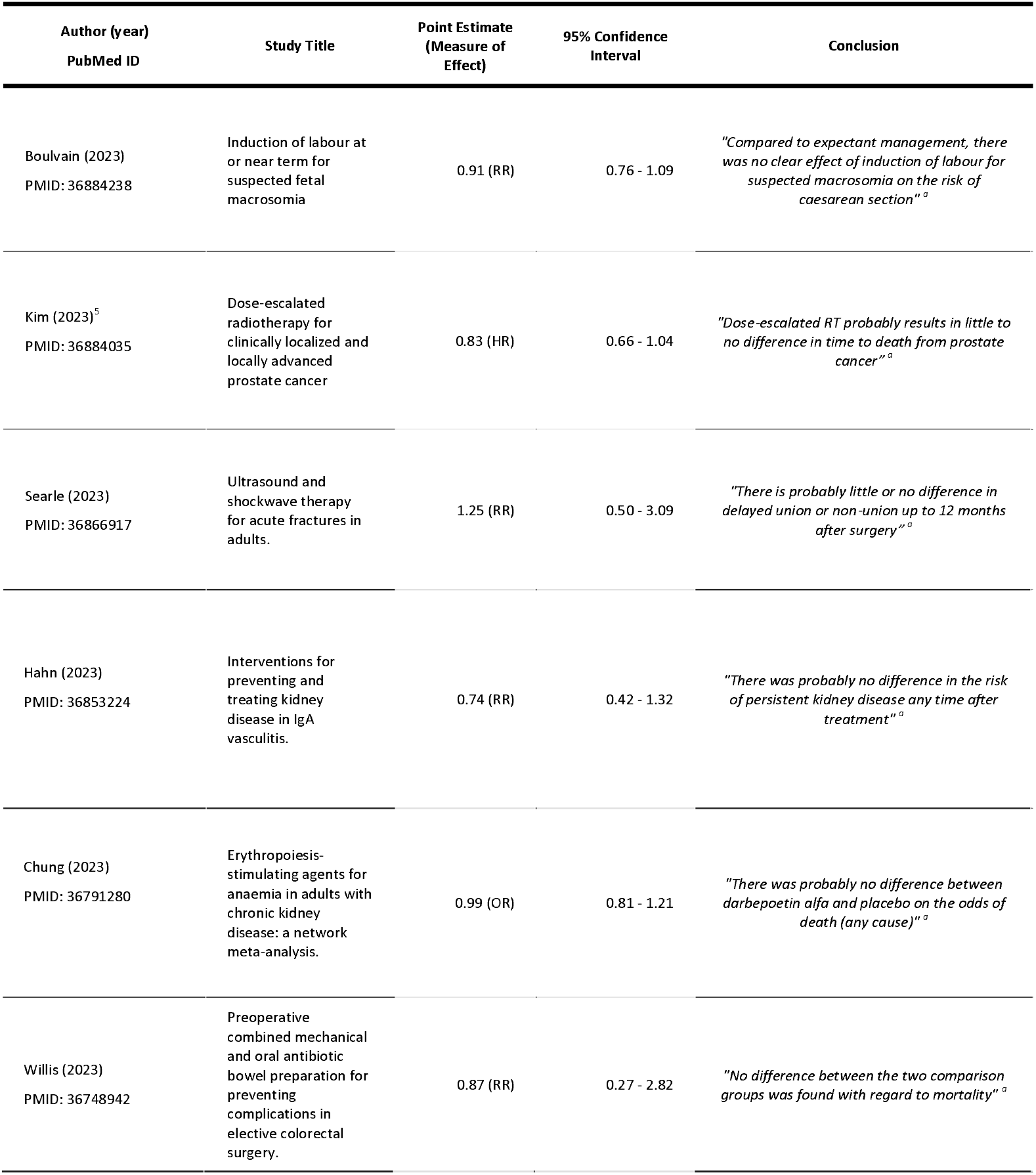
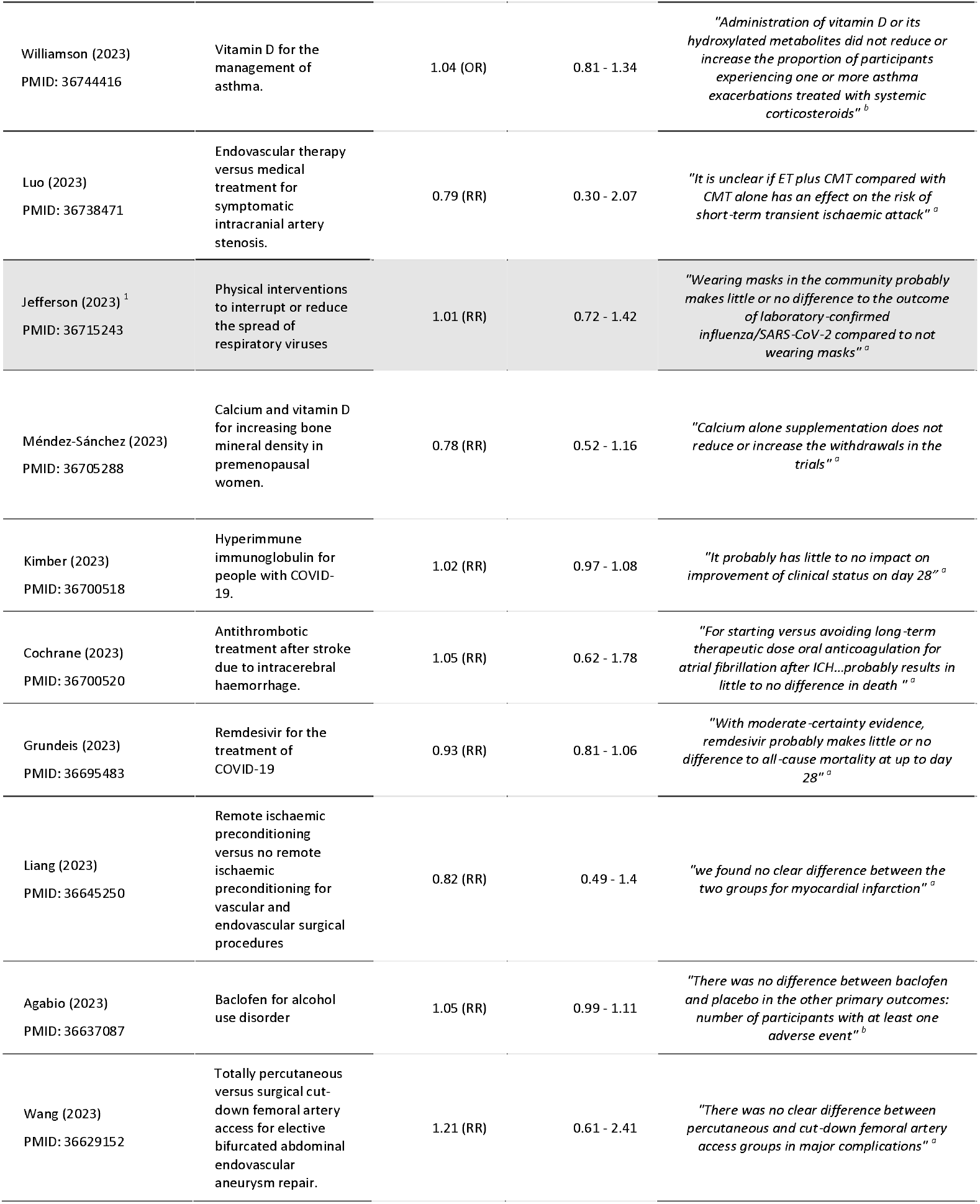
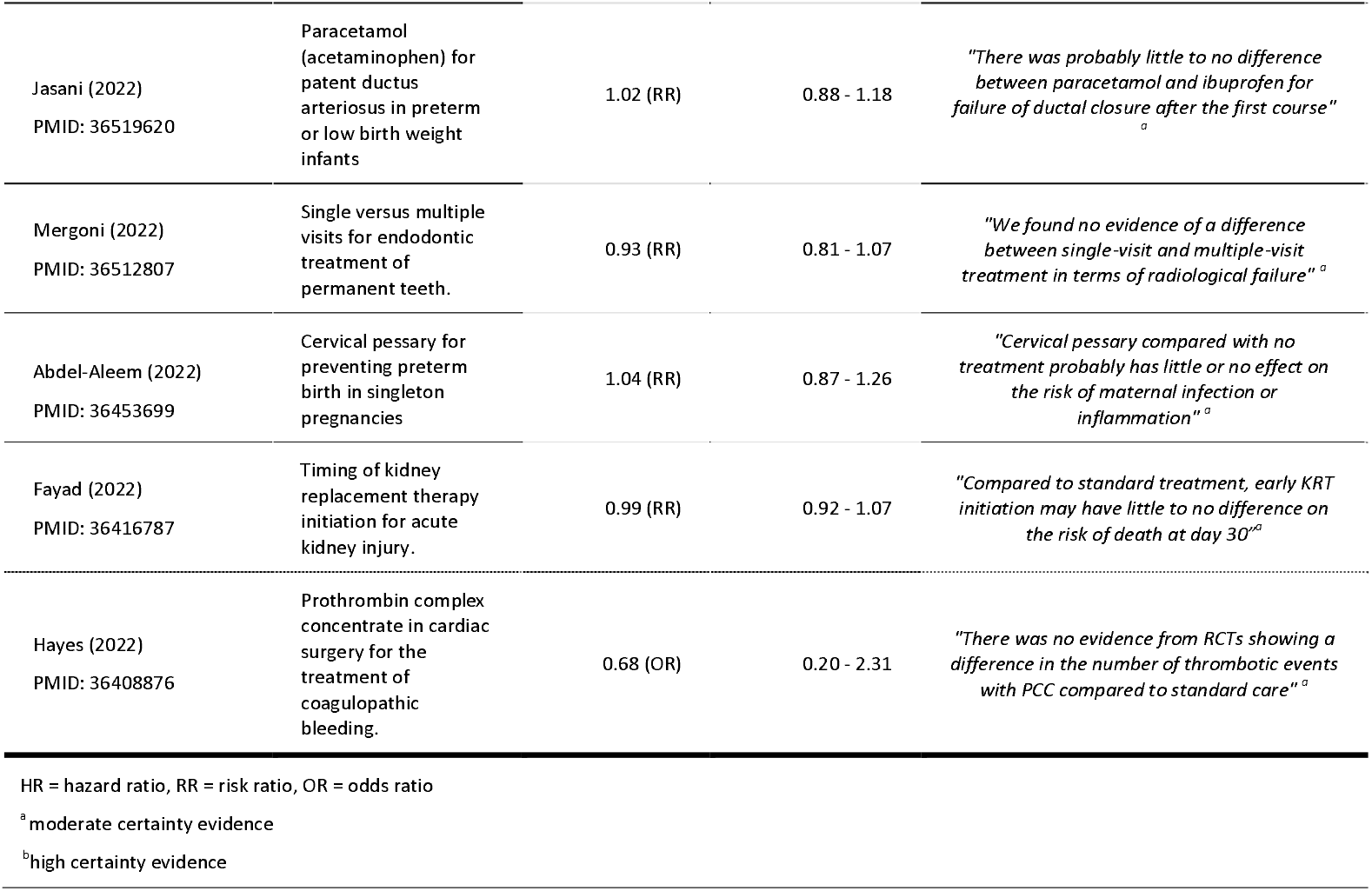
Results and conclusions from twenty consecutive Cochrane reviews reporting null findings published on or before March 9^th^, 2023

| Author (year)<br>PubMed ID | Study Title | Point Estimate<br>(Measure of<br>Effect) | 95% Confidence<br>Interval | Conclusion |
| --- | --- | --- | --- | --- |
| Boulvain (2023)<br>PMID: 36884238 | Induction of labour at or near term for suspected fetal macrosomia | 0.91 (RR) | 0.76 - 1.09 | *"Compared to expectant management, there was no clear effect of induction of labour for suspected macrosomia on the risk of caesarean section"*<sup>a</sup> |
| Kim (2023)<sup>5</sup><br>PMID: 36884035 | Dose-escalated radiotherapy for clinically localized and locally advanced prostate cancer | 0.83 (HR) | 0.66 - 1.04 | *"Dose-escalated RT probably results in little to no difference in time to death from prostate cancer"*<sup>a</sup> |
| Searle (2023)<br>PMID: 36866917 | Ultrasound and shockwave therapy for acute fractures in adults. | 1.25 (RR) | 0.50 - 3.09 | *"There is probably little or no difference in delayed union or non-union up to 12 months after surgery"*<sup>a</sup> |
| Hahn (2023)<br>PMID: 36853224 | Interventions for preventing and treating kidney disease in IgA vasculitis. | 0.74 (RR) | 0.42 - 1.32 | *"There was probably no difference in the risk of persistent kidney disease any time after treatment"*<sup>a</sup> |
| Chung (2023)<br>PMID: 36791280 | Erythropoiesis-stimulating agents for anaemia in adults with chronic kidney disease: a network meta-analysis. | 0.99 (OR) | 0.81 - 1.21 | *"There was probably no difference between darbepoetin alfa and placebo on the odds of death (any cause)"*<sup>a</sup> |
| Willis (2023)<br>PMID: 36748942 | Preoperative combined mechanical and oral antibiotic bowel preparation for preventing complications in elective colorectal surgery. | 0.87 (RR) | 0.27 - 2.82 | *"No difference between the two comparison groups was found with regard to mortality"*<sup>a</sup> |
| Williamson (2023)<br>PMID: 36744416 | Vitamin D for the management of asthma. | 1.04 (OR) | 0.81 - 1.34 | *"Administration of vitamin D or its hydroxylated metabolites did not reduce or increase the proportion of participants experiencing one or more asthma exacerbations treated with systemic corticosteroids"*<sup>b</sup> |
| Luo (2023)<br>PMID: 36738471 | Endovascular therapy versus medical treatment for symptomatic intracranial artery stenosis. | 0.79 (RR) | 0.30 - 2.07 | *"It is unclear if ET plus CMT compared with CMT alone has an effect on the risk of short-term transient ischaemic attack"*<sup>a</sup> |
| Jefferson (2023)<sup>1</sup><br>PMID: 36715243 | Physical interventions to interrupt or reduce the spread of respiratory viruses | 1.01 (RR) | 0.72 - 1.42 | *"Wearing masks in the community probably makes little or no difference to the outcome of laboratory-confirmed influenza/SARS-CoV-2 compared to not wearing masks"*<sup>a</sup> |
| Méndez-Sánchez (2023)<br>PMID: 36705288 | Calcium and vitamin D for increasing bone mineral density in premenopausal women. | 0.78 (RR) | 0.52 - 1.16 | *"Calcium alone supplementation does not reduce or increase the withdrawals in the trials"*<sup>a</sup> |
| Kimber (2023)<br>PMID: 36700518 | Hyperimmune immunoglobulin for people with COVID-19. | 1.02 (RR) | 0.97 - 1.08 | *"It probably has little to no impact on improvement of clinical status on day 28"*<sup>a</sup> |
| Cochrane (2023)<br>PMID: 36700520 | Antithrombotic treatment after stroke due to intracerebral haemorrhage. | 1.05 (RR) | 0.62 - 1.78 | *"For starting versus avoiding long-term therapeutic dose oral anticoagulation for atrial fibrillation after ICH...probably results in little to no difference in death"*<sup>a</sup> |
| Grundeis (2023)<br>PMID: 36695483 | Remdesivir for the treatment of COVID-19 | 0.93 (RR) | 0.81 - 1.06 | *"With moderate-certainty evidence, remdesivir probably makes little or no difference to all-cause mortality at up to day 28"*<sup>a</sup> |
| Liang (2023)<br>PMID: 36645250 | Remote ischaemic preconditioning versus no remote ischaemic preconditioning for vascular and endovascular surgical procedures | 0.82 (RR) | 0.49 - 1.4 | *"we found no clear difference between the two groups for myocardial infarction"*<sup>a</sup> |
| Agabio (2023)<br>PMID: 36637087 | Baclofen for alcohol use disorder | 1.05 (RR) | 0.99 - 1.11 | *"There was no difference between baclofen and placebo in the other primary outcomes: number of participants with at least one adverse event"*<sup>b</sup> |
| Wang (2023)<br>PMID: 36629152 | Totally percutaneous versus surgical cut-down femoral artery access for elective bifurcated abdominal endovascular aneurysm repair. | 1.21 (RR) | 0.61 - 2.41 | *"There was no clear difference between percutaneous and cut-down femoral artery access groups in major complications"*<sup>a</sup> |
| Jasani (2022)<br>PMID: 36519620 | Paracetamol (acetaminophen) for patent ductus arteriosus in preterm or low birth weight infants | 1.02 (RR) | 0.88 - 1.18 | *"There was probably little to no difference between paracetamol and ibuprofen for failure of ductal closure after the first course"*<sup>a</sup> |
| Mergoni (2022)<br>PMID: 36512807 | Single versus multiple visits for endodontic treatment of permanent teeth. | 0.93 (RR) | 0.81 - 1.07 | *"We found no evidence of a difference between single-visit and multiple-visit treatment in terms of radiological failure"*<sup>a</sup> |
| Abdel-Aleem (2022)<br>PMID: 36453699 | Cervical pessary for preventing preterm birth in singleton pregnancies | 1.04 (RR) | 0.87 - 1.26 | *"Cervical pessary compared with no treatment probably has little or no effect on the risk of maternal infection or inflammation"*<sup>a</sup> |
| Fayad (2022)<br>PMID: 36416787 | Timing of kidney replacement therapy initiation for acute kidney injury. | 0.99 (RR) | 0.92 - 1.07 | *"Compared to standard treatment, early KRT initiation may have little to no difference on the risk of death at day 30"*<sup>a</sup> |
| Hayes (2022)<br>PMID: 36408876 | Prothrombin complex concentrate in cardiac surgery for the treatment of coagulopathic bleeding. | 0.68 (OR) | 0.20 - 2.31 | *"There was no evidence from RCTs showing a difference in the number of thrombotic events with PCC compared to standard care"*<sup>a</sup> |

HR = hazard ratio, RR = risk ratio, OR = odds ratio
<sup>a</sup> moderate certainty evidence
<sup>b</sup> high certainty evidence

**Figure.**
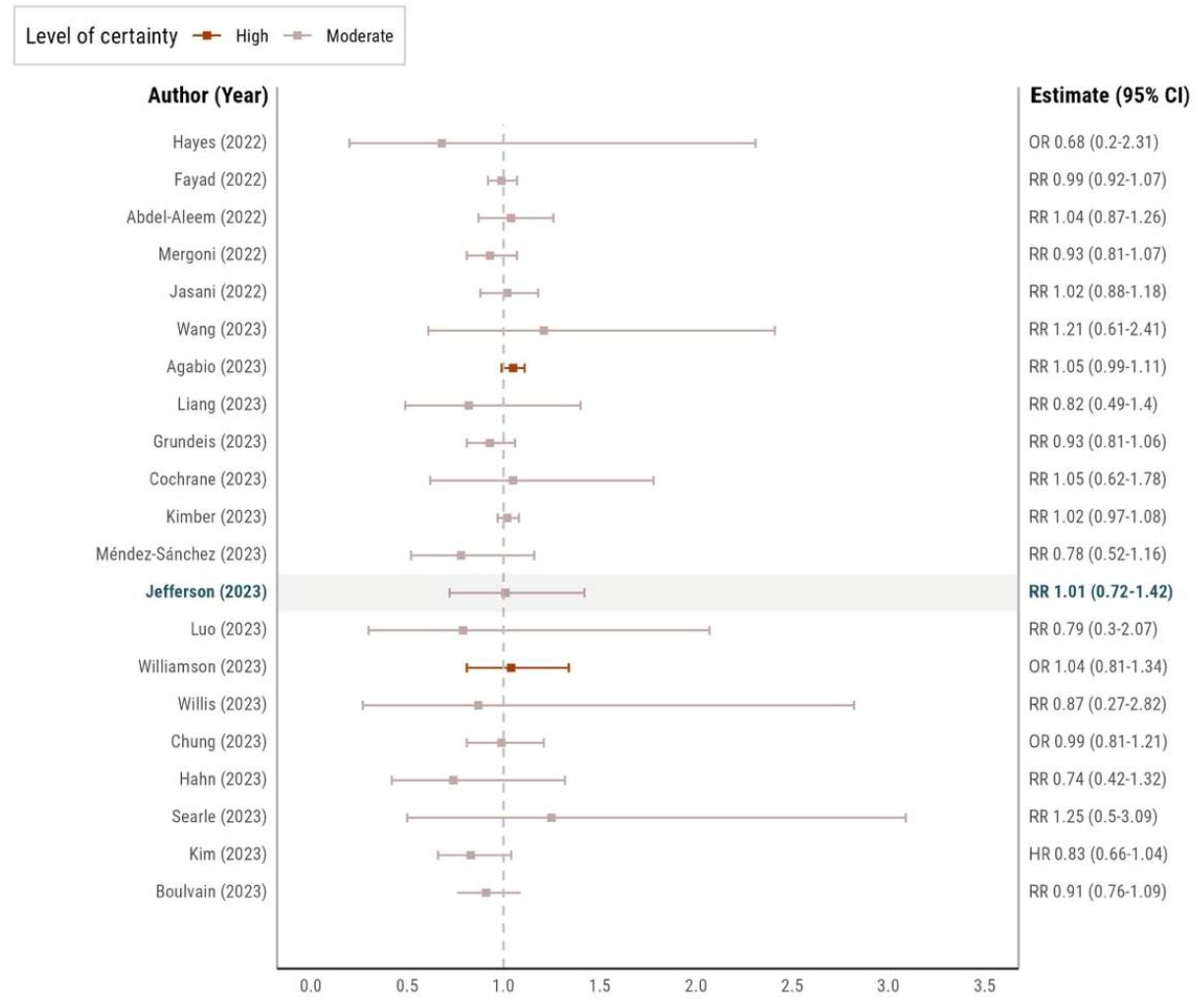
Effect estimates and 95% confidence intervals from selected Cochrane reviews. Forest plot showing point estimates and 95% confidence intervals from 20 consecutive Cochrane reviews with null findings published on or before March 9^th^, 2023. Point estimates are represented as risk ratios (RR), hazard ratios (HR), or odds ratios (OR). The dashed line indicates the line of no effect.

### Statistical analysis

Descriptive statistics were gathered. The primary outcome of interest was the specific reviewer language used in the interpretation of the findings. We concluded interpretations were consistent with Jefferson’s interpretations if they described the intervention as having no effect on the outcome, using the phrases “no difference” or “no effect” or “does not increase or decrease” when describing the effect of the intervention on the outcome. Interpretations describing the effect of the intervention on the outcome as being “unclear” were categorized as inconsistent with Jefferson’s language.

## Results

We found between November 21^st^, 2022, and March 9^th^, 2023, there were 20 Cochrane reviews that met the inclusion criteria. We found that 95% (n = 19) of the reviews used language that was consistent with Jefferson’s findings, while 5% (n = 1) described their results as “unclear”.

The argument that Jefferson’s findings could be compatible with some benefit would apply to all the analyses in the **Figure**. In some cases the confidence interval is primarily on the side of benefit, e.g. Kim et al. on the effect of dose-escalated radiotherapy on time to death from prostate cancer (HR 0.83, 95% CI 0.66 - 1.04).^5^ In comparison, Jasani’s finding on the effect of paracetamol compared with ibuprofen on failure of ductal closure (RR 1.02, 95% CI 0.88 - 1.18)^6^ and Jefferson’s study (RR 1.01, 95% CI 0.72 - 1.42)^1^ indicate no differences in outcome between the arms and have fairly balanced confidence intervals above and below one, indicating that they aren’t any more compatible with relative benefit than they are harm.

It’s worth noting that the precision around the estimates, represented by the widths of the confidence intervals, do vary between the studies. Agabio’s finding on the effect of baclofen on the number of participants with at least one adverse event in a study on alcohol use disorder^7^ has a relatively narrow confidence interval (RR 1.05, 95% CI 0.99 – 1.11) in comparison to Jefferson’s findings. It’s reasonable to be more skeptical of findings with wide confidence intervals and consider whether it is worth running additional high-quality studies to improve precision. However, null findings like these are still often enough to deprioritize an intervention or abandon it entirely, especially when those findings are derived from randomized trials.

## Discussion

We found that Jefferson’s conclusion on the effect of medical/surgical masks on laboratory-confirmed influenza or SARS-CoV-2 was similar to other reviews with null findings. For example, in their review assessing Remdesivir for the treatment of COVID-19^8^, Grundeis et al. conclude “*With moderate-certainty evidence, remdesivir probably makes little or no difference to all-cause mortality at up to day 28*”. The language used to describe the results is nearly identical to the language used by Jefferson et al. apart from the study-specific variables. In fact, it’s similar to most of the conclusions listed in the **Table**, with some variation depending on the certainty of the evidence. This is no coincidence; the Cochrane Handbook^9^ provides guidance on how to report and interpret findings and this is the language they recommend for reports where the size of the effect estimate is “trivial, small, unimportant effect, or no effect”, with the inclusion of the word “probably” for moderate certainty of evidence.

On these grounds, the conclusions made by Jefferson and colleagues were not only appropriate, but in line with the standardized approach created by Cochrane. Further, Jefferson’s comment in the media about there being “no evidence that they make any difference”^3^ is consistent with their conclusion in the Cochrane review in which they stated, “Wearing masks in the community probably makes little or no difference to the outcome of laboratory-confirmed influenza/SARS-CoV-2 compared to not wearing masks.”^1^

The pooling of the best available evidence in meta-analyses informs our understanding of how well interventions work. Null findings often do contribute to decision-making, despite the fact that they don’t provide conclusive proof that an intervention doesn’t work under any circumstances. We found no obvious difference between Jefferson’s review and other recent reviews that would justify the differential interpretation and treatment of this study or the unprecedented comments made over its findings. Clarifying comments of the editor-in-chief of Cochrane appear unjustified.

## Data Availability

All data produced in the present work are contained in the manuscript

